# Novel syndromic neurodevelopmental disorder caused by de novo deletion of *CHASERR*, a long noncoding RNA

**DOI:** 10.1101/2024.01.31.24301497

**Authors:** Vijay S. Ganesh, Kevin Riquin, Nicolas Chatron, Kay-Marie Lamar, Miriam C. Aziz, Pauline Monin, Melanie O’Leary, Julia K. Goodrich, Kiran V. Garimella, Eleina England, Esther Yoon, Ben Weisburd, Francois Aguet, Carlos A. Bacino, David R. Murdock, Hongzheng Dai, Jill A. Rosenfeld, Lisa T. Emrick, Shamika Ketkar, Undiagnosed Diseases Network, Yael Sarusi, Damien Sanlaville, Saima Kayani, Brian Broadbent, Bertrand Isidor, Alisée Pengam, Benjamin Cogné, Daniel G. MacArthur, Igor Ulitsky, Gemma L. Carvill, Anne O’Donnell-Luria

**Author notes:** Corresponding authors: Anne O’Donnell-Luria,; Gemma L. Carvill. These authors contributed equally to this work (Vijay S. Ganesh, Kevin Riquin).

## Abstract

Genes encoding long non-coding RNAs (lncRNAs) comprise a large fraction of the human genome, yet haploinsufficiency of a lncRNA has not been shown to cause a Mendelian disease. *CHASERR* is a highly conserved human lncRNA adjacent to *CHD2–*a coding gene in which *de novo* loss-of-function variants cause developmental and epileptic encephalopathy. Here we report three unrelated individuals each harboring an ultra-rare heterozygous *de novo* deletion in the *CHASERR* locus. We report similarities in severe developmental delay, facial dysmorphisms, and cerebral dysmyelination in these individuals, distinguishing them from the phenotypic spectrum of *CHD2* haploinsufficiency. We demonstrate reduced *CHASERR* mRNA expression and corresponding increased *CHD2* mRNA and protein in whole blood and patient-derived cell lines–specifically increased expression of the *CHD2* allele in *cis* with the *CHASERR* deletion, as predicted from a prior mouse model of *Chaserr* haploinsufficiency. We show for the first time that *de novo* structural variants facilitated by Alu-mediated non-allelic homologous recombination led to deletion of a non-coding element (the lncRNA *CHASERR*) to cause a rare syndromic neurodevelopmental disorder. We also demonstrate that *CHD2* has bidirectional dosage sensitivity in human disease. This work highlights the need to carefully evaluate other lncRNAs, particularly those upstream of genes associated with Mendelian disorders.

## Introduction

Developmental and epileptic encephalopathies are genetically and phenotypically heterogeneous disorders causing severe global neurodevelopmental delay^1^. *De novo* loss-of-function variants in several genes, including chromodomain helicase DNA-binding protein 2 (*CHD2*), are known to cause early-onset epileptic encephalopathy^2^ (MIM 615369). CHD2 is a DNA-binding protein implicated in chromatin remodeling, with a role in neurogenesis of cortical neurons and interneurons^3–6^. The most common phenotypes in *CHD2*-related neurodevelopmental disorders include seizures in 96% of individuals, with a median age of onset of 30 months, developmental delay and intellectual disability in 95%, and autism spectrum disorder in 56%^7^. Brain imaging in these patients is often normal, or with generalized atrophy in some individuals^8^. To date, all pathogenic variants in *CHD2* are *de novo*, with a suspected loss-of-function mechanism; the majority are truncating variants or deletions, and a few are missense variants in the highly conserved DNA-binding or helicase domains^7^. Consistent with the loss of function constraint seen for *CHD2* in gnomAD^9^ (pLI 1, LOEUF 0.17), the Clinical Genome Resource (ClinGen) curated *CHD2* as a haploinsufficient gene. There is recent evidence from analysis of rare copy number variation in the human population to support triplosensitivity of *CHD2*^*10*^, but increased *CHD2* expression has not yet been demonstrated in a neurodevelopmental disorder (NDD).

*CHASERR* is a highly-conserved long non-coding RNA (lncRNA) adjacent to *CHD2*. Its mouse homolog (*Chaserr*) mediates *cis*-acting transcriptional repression of *Chd2* and is essential for postnatal survival^11^. In the nuclei of mouse embryonic stem cells, *Chaserr* RNA localizes within a topologically associated domain (TAD) containing the *Chaserr* gene and Chd2 protein^12^.

However, to date, *CHASERR* has not been implicated in human disease. Only a single lncRNA has been associated with a human Mendelian disease–homozygous deletions of *LINC01956* causing a severe congenital limb and hindbrain malformation syndrome^13^. Haploinsufficiency of a lncRNA, particularly via increased target gene expression, has not been previously associated with a human Mendelian condition.

Here we report three unrelated individuals with a syndromic, early-onset NDD each harboring a heterozygous *de novo* deletion involving *CHASERR* that leaves the promoter and coding region of *CHD2* intact. We report their phenotypic similarities of developmental delay, shared facial dysmorphisms, and cerebral dysmyelination–all with a more severe phenotype compared to the known clinical spectrum of *CHD2* haploinsufficiency. In two affected individuals we demonstrate 50% reduction of *CHASERR* transcript and corresponding overexpression of *CHD2*, specifically of the *CHD2* allele in *cis* with the *de novo CHASERR* deletion. This is the first report of *de novo* heterozygous deletions involving a lncRNA causing a Mendelian disease, providing evidence of bidirectional dosage sensitivity of *CHD2* in human brain development, and highlighting the need to more closely investigate highly-conserved lncRNAs in undiagnosed diseases.

## Methods

### Clinical data and specimen collection

Individual 1 was enrolled in the Undiagnosed Diseases Network, with informed consent obtained for patient and parental enrollment according to an institutional review board protocol (IRB) at the National Institutes of Health (#15HG0130). Individual 1, sibling, and parents were also enrolled in the Rare Genomes Project at the Broad Institute of MIT and Harvard through an IRB approved by Mass General Brigham (Protocol# 2016P001422).

Informed consent for Individuals 2 and 3 was obtained from the legal guardians according to the French law on Bioethics and following the Helsinki Declaration, as part of a study approved by the CHU de Nantes Ethics Committee (Research Programme “Génétique Médicale” DC-2011-1399).

The MatchMaker Exchange (MME)^14^ network facilitated connection between the investigators for Individuals 1 and 2, through the GeneMatcher node^15^.

### Sequencing and genomic analysis

Genomic DNA was isolated from peripheral blood, and trio genome sequencing (mother, father, and proband) was performed on samples obtained from Families 1-3. Long read genome sequencing was additionally performed on Family 1 using the Pacific Biosciences (PacBio) circular consensus sequencing (CCS) protocol. Additional details regarding genomic analyses are provided in the Supplementary Appendix.

### Expression analysis

RNA-sequencing was performed on whole blood and cultured fibroblasts on Individuals 1 and 2, and compared to corresponding controls from the GTEx Consortium^16^. Additionally, RNA-seq was performed on induced pluripotent stem cells (iPSCs) and cultured neural precursor cells (NPCs) from Individual 1, and protein quantification of CHD2 performed on iPSCs from Individual 1. Details regarding expression/abundance analyses are provided in the Supplementary Appendix.

## Results

### Clinical evaluations

#### Individual 1

Individual 1 is a female born to non-consanguineous parents with birth weight 3.4 kg (60th percentile), length 47 cm (15th percentile), and head circumference 35 cm (75th percentile) (Table 1). Initial examination was notable for generalized hypotonia and facial dysmorphisms including widely spaced eyes with upslanting palpebral fissures, anteverted nares, long philtrum, low set ears, and mild micrognathia.

**Table 1:**

| **Clinical features of CHASERR vs. CHD2 haploinsufficiency** |  |  |  |  |
| --- | --- | --- | --- | --- |
|  | Individual 1 | Individual 2 | Individual 3 | Individuals with CHD2 haploinsufficiency |
| **Measurements** |  |  |  |  |
| **Weight (at birth)** | 3.4 kg (60th percentile) | 4.0 kg (90th percentile) | 3.6 kg (75th percentile) | Normal |
| **Length (at birth)** | 47 cm (15th percentile) | 54 cm (90th percentile) | 50.5 cm (62th percentile) | Normal |
| **Head circumference (at birth)** | 35 cm (75th percentile) | 36 cm (50th percentile) | 36.8 cm (93th percentile) | Normal |
| **Weight (age 2 yrs)** | 10.1 kg (17th percentile) | 13.4 kg (70th percentile) | 9.8 kg (3rd percentile) | Normal |
| **Height (age 2 yrs)** | 74.5 cm (<1st percentile) | 95 cm (70th percentile) | 81 cm (2nd percentile) | Normal |
| **Head circumference (age 2 yrs)** | 44.8 cm (1st percentile) | 47.5 cm (30th percentile) | 45 cm (1st percentile) | Normal |
| **Tone at birth** | Hypotonia | Hypotonia | Normal | Normal |
| **Facial dysmorphisms** | Yes | Yes | Yes | Yes |
| **Electrographic and MRI Features** |  |  |  |  |
| **First EEG (age 1-5 years)** | Normal | Normal | Abnormal (posterior slowing, no epileptiform discharges) | Abnormal |
| **Latest EEG** | Polymorphic, rhythmic delta slowing (2 Hz), generalized polyspike and wave (GSPW) discharges | Symmetric, continuous theta-delta slowing, without epileptiform discharges | Symmetric, polymorphic, rhythmic slowing, with spike-and-wave discharges | - |
| **Medically refractory epilepsy** | No | No | No | Yes |
| **Seizure type(s)** | Infantile spasms, myoclonic | None | Infantile spasms, tonic | Absence, atonic, myoclonic, tonic |
| **Generalized convulsions** | No | No | No | Yes (>95%) |
| **Photosensitivity** | No | N/A | N/A | Yes |
| **Brain MRI** | cortical atrophy, cerebral hypomyelination, thin corpus callosum | cortical atrophy, cerebral hypomyelination, thin corpus callosum | cortical atrophy, cerebral hypomyelination, thin corpus callosum | no cerebral dysmyelination, cortical atrophy rare |
| **Milestones and Functional Status** |  |  |  |  |
| **Global developmental delay** | Yes, severe | Yes, severe | Yes, severe | Yes, variable |
| **Intellectual disability** | Yes, severe | Yes, severe | Yes, severe | Yes, variable |
| **Age at first neurodevelopmental symptoms/delay** | 1 month | 1 month | 1 month | 24 months (median) |
| **Able to communicate with words** | No | No | No | 90% able to |
| **Able to walk independently** | No | No | No | 88% able to |
| **Able to grasp small objects** | No | No | No | 85% able to |
| **Able to dress self** | No | No | No | 50% able to |
| **Able to use spoon/fork** | No | No | No | 75% able to |
| **Able to drink from cup** | No | No | No | 75% able to |

Brain MRI in the infantile period demonstrated mild hypomyelination with hypoplastic optic nerves, optic chiasm, and optic tracts. A normal electroencephalogram (EEG) was noted at this time. As a toddler she was diagnosed with hypsarrhythmia and infantile spasms, with an abnormal interictal EEG marked by polyspike and wave discharges without discrete electrographic or clinical seizures. Multiple anticonvulsants were started (levetiracetam, clobazam, vigabatrin) with some clinical improvement in spasms without improvement in the electrographic abnormalities. CSF neurotransmitter and amino acid evaluation was normal. Urine sialic acid (Greenwood Genetics), urine mucopolysaccharides screen (Mayo Clinic), serum lysosomal enzyme testing (Lysosomal Diseases Testing Laboratory), and congenital disorders of glycosylation testing (Mayo Clinic) were all normal. SNP-based chromosomal microarray (Baylor Genetics), clinical trio exome sequencing (Ambry Genetics), and genome-wide DNA methylation episignature assay (EpiSign, Greenwood Genetics) were all normal/negative. Repeat brain MRI as a toddler showed generalized cortical atrophy and persistence of global dysmyelination with thinning of the corpus callosum and reduced brainstem volume, without evidence of focal cortical dysplasias or abnormal gyrification, and no ischemia or hemorrhage (Figure 1), and Lennox-Gastaut syndrome was diagnosed following a repeat video EEG, still without any discrete clinical seizures.

**Figure 1:**
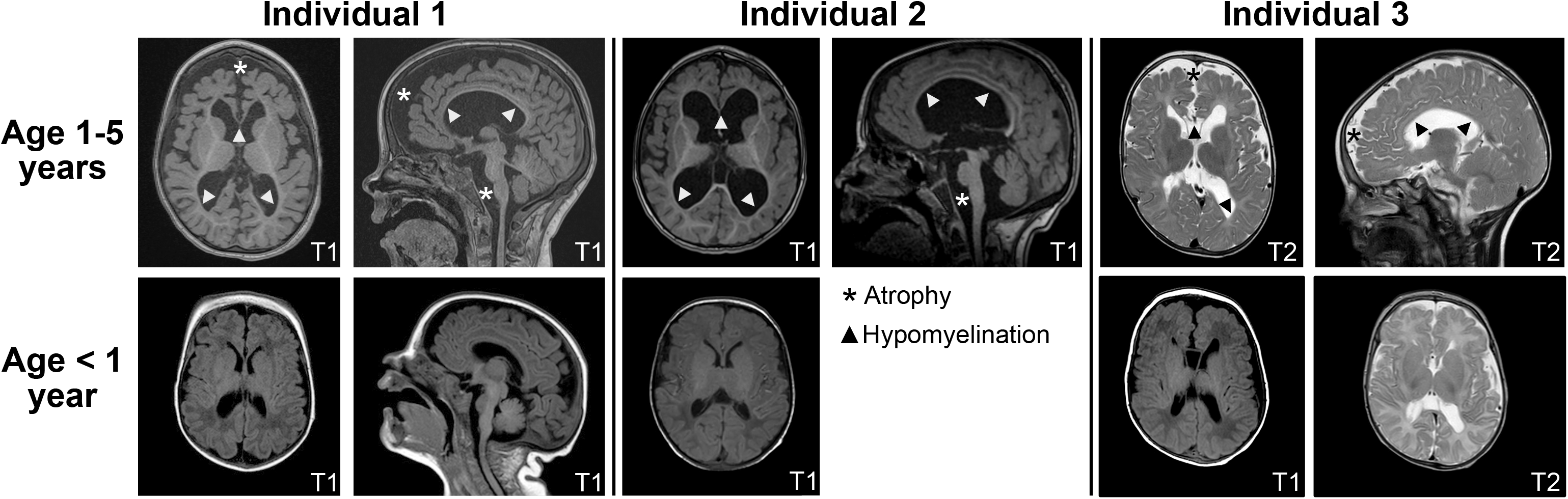
Brain MRI in individuals with *CHASERR* haploinsufficiency. Brain magnetic resonance imaging (MRI) demonstrates frontal-predominant cortical atrophy with reduced volume of the brainstem (asterisks), and generalized hypomyelination of the corpus callosum and subcortical white matter (arrowheads) (top row), all less apparent in the first year of life (bottom row). T1, T2 = MRI sequence protocols.

#### Individual 2

Individual 2 is a male born to non-consanguineous parents after induction of labor at full term with a birth weight of 4.0 kg (90th percentile), length 54 cm (90th percentile), and head circumference 36 cm (50th percentile) (Table 1). Pregnancy was uneventful. Feeding difficulties, bilateral adducted thumbs, and poor weight gain were noted in the infantile period, with similar facial dysmorphisms as Individual 1, including widely spaced eyes, anteverted nares, and long philtrum, and additionally mild hypertrichosis with thick eyebrows. Neurologic exam showed axial hypotonia and peripheral hypertonia with brisk reflexes, and bilateral optic atrophy. As a toddler he still could not yet sit or speak words and had dystonic movements in the bilateral arms and legs. Brain MRI in the infantile period was normal, but repeat MRI as a toddler showed global dysmyelination with thinning of the corpus callosum, reduced brainstem volume, and cortical atrophy (Figure 1), withEEG showing symmetric continuous theta-delta slowing, with intact sleep architecture, without definite epileptiform discharges. No anticonvulsant treatments have been started and he is treated with trihexyphenidyl with incomplete resolution of dystonic movements.

#### Individual 3

Individual 3 is a male born to non-consanguineous parents at full term after an uneventful pregnancy, with a birth weight of 3.64 kg (75th percentile), length 50.5 cm (60th percentile) and head circumference 36.8 cm (90th percentile) (Table 1). Neonatal examination showed cryptorchidism and distal arthrogryposis of the limbs with slightly proximally placed thumbs.

Atypical spasms with desaturations on pulse oximetry were noted in the infantile period. Initial EEG showed a slow background rhythm in posterior regions, and polymorphic right temporal-occipital slowing, without electrographic seizures. Video EEG performed as a toddler showed persistence of slow, disorganized background without obvious asymmetry, with rare spike-and-wave discharges occurring mostly during sleep. A long-term EEG study later in childhood captured epileptic seizures with electrographic-clinical correlate of extension of the upper limbs and flexion of the lower limbs. Levetiracetam and vigabatrin did not significantly improve the paroxysmal spasms or EEG background. Brain MRI performed shortly after birth showed cortical hypomyelination with thinning of the corpus callosum, more apparent on repeat MRI as an infant (Figure 1). As a toddler there is severe global developmental delay with preserved tracking and regarding, expressive language limited to vocalizations, and inability to sit or roll over.

### *Detection and phasing of de novo deletions in* CHASERR

In Individuals 1-3, short read genome sequencing identified 22 kb, 8.4 kb, and 25 kb heterozygous *de novo* deletions, respectively, that each overlap the promoter and first three exons of *CHASERR* (Figure 2A), without deleting other nearby coding genes including *CHD2* or its promoter.

**Figure 2:**
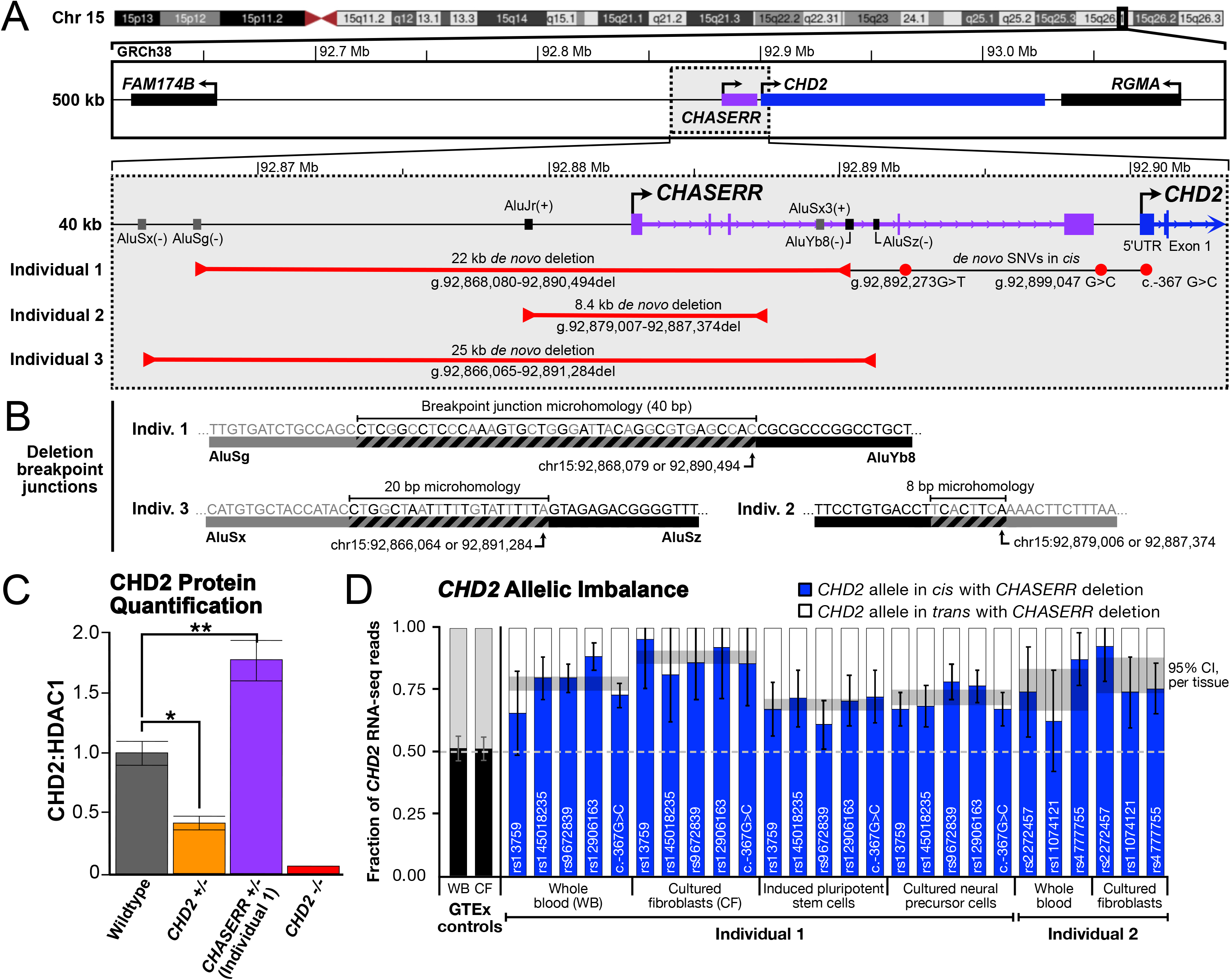
*De novo CHASERR* deletions and characterization of *CHD2* expression. **A**. Human chromosome 15q26.1, with a 500 kb inset showing a gene-sparse region upstream of the long non-coding RNA gene *CHASERR* (purple) and its tandem coding gene *CHD2* (blue). Arrows = direction of transcription. *De novo* deletions (red bracketed lines) in Individual 1 (22 kb), Individual 2 (8.4 kb), and Individual 3 (25 kb) overlap the promoter and first three exons of *CHASERR*. Long read DNA sequencing in Individual 1 identified three *de novo* single nucleotide variants (SNVs, red dots) in *cis* with the *CHASERR* deletion. **B**. Breakpoint mapping for the deletions in Individuals 1 and 3 shows 40 bp and 20 bp microhomology, respectively, at the junction between two negative-strand oriented SINE/Alu elements (AluSg and AluYb8 for Indiv. 1; AluSx and AluSz for Indiv. 3). Short read genome sequencing in Individual 2 maps the breakpoint near two positive-strand oriented SINE/Alu elements (AluJr, AluSx3), with 8 bp microhomology at the junction. **C**. Increased CHD2 protein abundance in patient-derived neural precursor cells from Individual 1 (purple) compared to iPSCs from unaffected controls (gray), an individual with *CHD2* haploinsufficiency (orange), and *CHD2*-/- iPSCs (red). CHD2 abundance in haploinsufficient control iPSC lines was 50% relative to wildtype controls. Error bars = ± standard error. ^*^ p=0.0009, ^**^ p=0.0024 (t-test). **D**. RNA-seq from whole blood (WB), cultured fibroblasts (CF), cultured neural precursor cells, and induced pluripotent stem cells from Individual 1, and cultured fibroblasts and whole blood from Individual 2, all show allelic imbalance towards expression of the *CHD2* allele in *cis* with the *CHASERR* deletion. Control whole blood and cultured fibroblasts from GTEx Consortium participants (black bars) show no allelic imbalance in *CHD2*. Error bars = 95% confidence interval (assuming binomial distribution).

Long read DNA sequencing in Individual 1 confirmed the *de novo* 22 kb deletion and delineated the breakpoint to two negative strand-oriented SINE/Alu elements (AluSg, AluYb8) with 40 bp of microhomology at the breakpoint junction (Figure 2B), suggesting a mechanism of replicative non-homologous double-stranded DNA break repair. In addition, long read sequencing confirmed three non-coding *de novo* SNVs *in cis* with the 22 kb deletion (Figure 2A), likely generated by error-prone polymerase during the break repair process^17^. The *CHASERR* deletion is most likely mediating the phenotype, not the incidentally identified SNVs, as these noncoding variants are not at highly conserved positions and no *de novo* SNVs were identified in Individuals 2 or 3 in the *CHASERR* locus.

Short read genome sequencing data in Individuals 2 and 3 showed a similar deletion mechanism, with breakpoints within two negative-strand oriented SINE/Alu elements (AluSx, AluSz) with 20 bp of microhomology at the junction in Individual 3, and in Individual 2 a deletion in close proximity to two positive stand-oriented SINE/Alu elements (AluJr, AluSx3) with 8 bp of microhomology at the junction (Figure 2B). The breakpoint junction microhomology sequences in Individuals 1 and 3 map to a known recombination hotspot within SINE/Alu elements^18^.

In Individual 1, phasing of single nucleotide variants (SNVs) by long read sequencing established that the *de novo* deletion in *CHASERR* occurred on the paternally-inherited chromosome. In Individual 2, the *de novo* deletion also occurred on the paternally-inherited chromosome, with phasing ascertained by the detection of a Mendelian violation at an SNV within the deletion interval (g.92882694C>T). Parent of origin for the *de novo* deletion in *CHASERR* in Individual 3 was not able to be determined from short read genome sequencing.

A genome-wide search for other *de novo* variants in the three trios identified a likely pathogenic variant for Individual 1 in *CITED2* (NM_006079.5, c.701A>C, p.Glu234Ala) that was suspected to account for Individual 1’s atrial septal defect but not the neurologic phenotype. For Individuals 1-3, standard variant analysis using both dominant and recessive models did not identify any other pathogenic or likely pathogenic SNVs, indels, short tandem repeat expansions, or structural variants in the nuclear or mitochondrial genomes.

### CHD2 *mRNA expression, allelic imbalance, and protein abundance*

RNA sequencing (RNA-seq) of whole blood from Individual 1 demonstrated haploinsufficiency of *CHASERR* compared to an unaffected sibling, with 6.95 transcripts per million (TPM) in Individual 1 compared to 23.92 TPM in the unaffected sibling. Whole blood RNA-seq expression of *CHD2* (the tandem gene downstream to *CHASERR*) showed increased expression in the proband (23.9 TPM) relative to the unaffected sibling (18.17 TPM) and adult GTEx samples (median TPM 10.95), as expected from the *Chaserr+/-* mouse model^11^. No other transcriptome-wide gene expression outliers were seen in whole blood compared to GTEx controls (Supplementary Figure S1). We observed a statistically significant 1.8 fold increase in CHD2 protein abundance in iPSCs from Individual 1, supporting the hypothesis that *CHASERR* deletion leads to an increase in *CHD2* mRNA expression and protein abundance (Figure 2C, Supplementary Figure S2; p<0.0024, t-test).

On closer inspection of *CHD2* expression from RNA-seq data, consistent allelic imbalance was noted in all available tissues, including whole blood, cultured fibroblasts, iPSCs, and cultured neural precursor cells (NPCs) from Individual 1, and whole blood and cultured fibroblasts from Individual 2 (Figure 2D), across all phaseable SNVs in *CHD2*. Moreover, the imbalance was skewed towards the allele in *cis* with the *CHASERR* deletion (i.e. the paternal allele), suggesting cis-regulatory derepression of *CHD2* expression, as expected from the *Chaserr*+/- mouse model^11^. The allelic imbalance accounts for the mildly increased (i.e. derepressed) total *CHD2* expression. Collectively, these results demonstrate that *CHASERR* is a negative *cis* regulator of *CHD2*, and that *CHASERR* deletion results in increased CHD2 abundance.

## Discussion

We report the first case series of *de novo* heterozygous deletions of a lncRNA and resultant increased target gene (*CHD2*) expression causing a human neurodevelopmental disorder, with three unrelated individuals sharing a syndrome of severe encephalopathy with cerebral hypomyelination and dysmorphic facies. In all three individuals, the deletions overlap *CHASERR* without including its downstream tandem gene, *CHD2*. This results in a 50% reduction in *CHASERR* transcript expression with a corresponding increase in CHD2 mRNA expression and protein abundance, specifically due to upregulation of the *CHD2* allele in *cis* with the *CHASERR* deletion. This functional effect matches what was observed in mice, where *Chaserr* haploinsufficiency causes *Chd2* overexpression in *cis*^*11*^.

All three individuals with *CHASERR* deletions are more impaired than even the most severe end of the *CHD2* haploinsufficiency phenotypic spectrum. Whereas most individuals with *CHD2* haploinsufficiency are verbal and ambulatory, none of the individuals reported here achieved those developmental milestones. All three individuals also have a different electrographic phenotype (none or few clinical seizures, no photosensitivity) and radiographic phenotype (cerebral dysmyelination, Figure 1) than what is typically seen in *CHD2*-related disorders (Table 1), altogether suggesting that *CHASERR* haploinsufficiency represents a new disease clinically distinguishable from *CHD2* haploinsufficiency-related disease.

Since the *de novo* deletions of *CHASERR* reported here implicate Alu-mediated non-allelic homologous recombination as the causal mechanism, we anticipate reanalysis of genome sequencing from other large cohorts of neurodevelopmental disorders (or array data when there is sufficient probe coverage of *CHASERR*) to uncover additional cases of *CHASERR* haploinsufficiency.

Our results extend the model of dosage sensitivity of *CHD2* in human brain development beyond haploinsufficiency alone, by strongly suggesting that *CHD2* overexpression via loss of *CHASERR* also perturbs normal human brain development. We are not aware of *CHD2*-only duplications (i.e. duplications dissociating *CHD2* from *CHASERR*) in either cases or controls. *CHD2* triplosensitivity has been recently inferred from gene dosage-sensitivity metrics imputed from the largest study of rare copy number variation (*CHD2* pTriplo = 0.986, with the defined threshold for a strong effect > 0.94)^10^. *CHD2* haploinsufficiency is reported to disrupt human cortical interneuron development via loss of binding to enhancer sites for multiple transcription factors at specific timepoints to orchestrate the switch from neural progenitor proliferation to neuronal differentiation^6,19^. It is possible that exquisite sensitivity to the timing of this switch may account for the bidirectional dosage sensitivity of *CHD2. CHD8*, a related chromatin-modifying gene that also causes human NDD, also has evidence to suggest dosage sensitivity to the maintenance of pluripotency and neural differentiation in a murine model^20^. Collectively these results suggest that chromatin remodelers and other genes that modulate gene expression are particularly sensitive to gene dosage, including *MECP2* and *SETBP1*, and indeed many of these genes are bidirectionally sensitive in genome-wide analysis^10,21,22^.

This study and prior research in *Chaserr* haploinsufficient mice^11^ suggest that *CHASERR* may be an appropriate RNA-therapeutic target to rescue *CHD2* haploinsufficiency. However, caution is needed as excessive *CHD2* is also associated with poor, and potentially worse, neurological outcomes. This “Goldilocks problem” will need to be addressed, potentially through preclinical models of partial inhibition of *CHASERR*.

Overall, our findings highlight the essential role of a highly conserved lncRNA in human brain development and disease. *Chaserr*-mediated regulation of *Chd2* is suggested to be part of an autoregulatory feedback loop^23^, highlighting that *Chaserr* and related lncRNAs are adapted to regulate target genes that require exquisite spatial and temporal expression. As transcripts that do not need to function in the cytoplasm, such lncRNAs are potentially well-suited to sense the activity of chromatin-related proteins and then fine-tune gene expression in the nucleus. As such, it is expected that genetic lesions in lncRNA that regulate genes in *cis* will lead to imbalances in expression of these tandem genes, potentially resulting in disease. Yet lncRNA are largely overlooked in clinical genomic analyses. Indeed, homozygous deletions of the lncRNA *LINC01956* were recently shown to cause a severe congenital limb and brain malformation syndrome via disruption of *cis-*acting regulation of the nearby gene *EN1*^*13*^, similar to the tandem gene structure and regulatory model of *CHASERR-CHD2*. Altogether this highlights the emerging role for lncRNAs in human Mendelian conditions, and suggests that *de novo* SVs that encompass lncRNAs be examined closely in individuals with undiagnosed disorders.

## Data Availability

All data produced in the present study are available upon reasonable request to the authors.

