## Supplementary Appendix for "Novel syndromic neurodevelopmental disorder caused by de novo deletion of *CHASERR*, a long noncoding RNA"

**Table of Contents**


1. Supplementary Figures
    a. Supplementary Figure S1
    b. Supplementary Figure S2
2. Supplementary Methods
    a. Short-read genome sequencing
    b. Long-read genome sequencing
    c. Patient-derived cell lines
    d. RNA and cDNA amplicon sequencing
        i. Individual 1
        ii. Individual 2
    e. Immunoblot for CHD2 abundance
3. Supplementary References

*Supplementary Figure S1: RNA-seq outlier expression analysis for Individual 1*

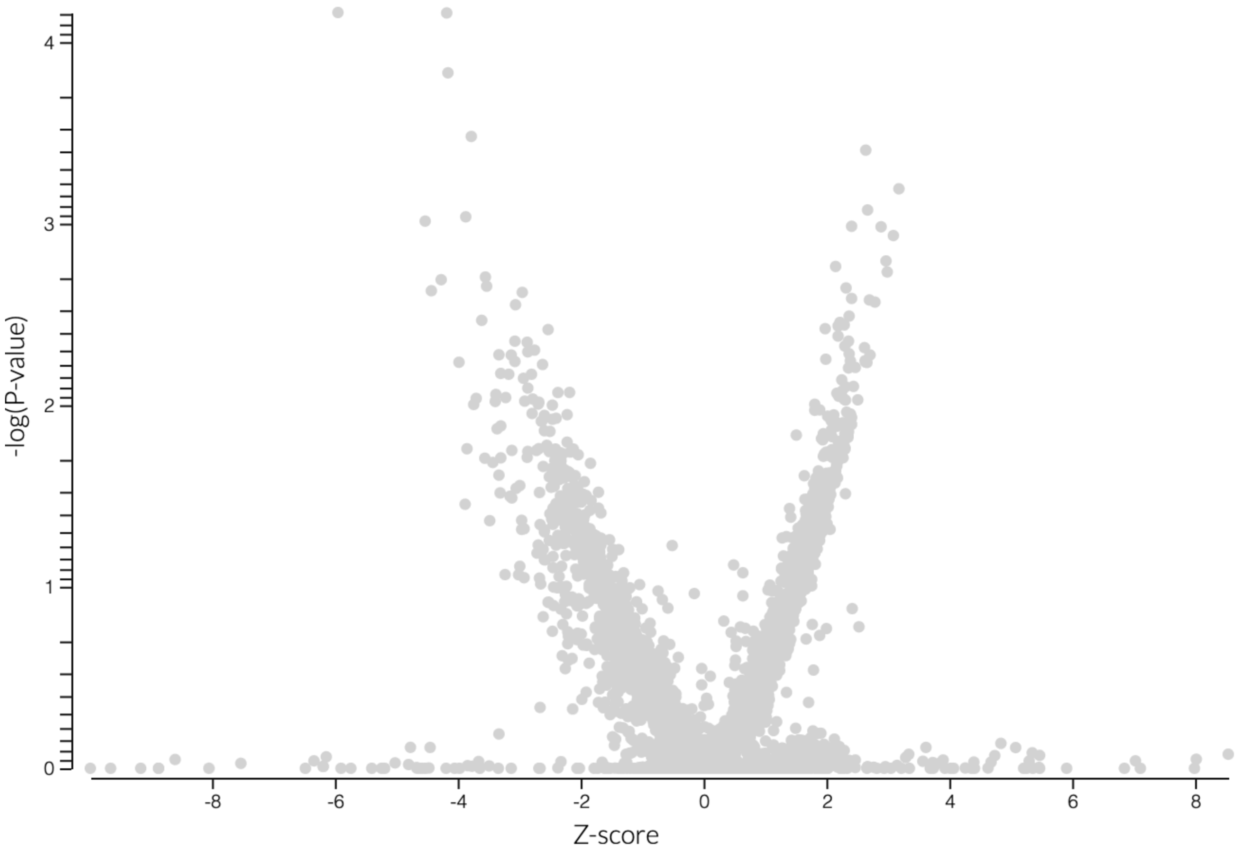


*Supplementary Figure S1 Legend:*

Volcano plot of OUTRIDER analysis of whole blood RNA-seq from Individual 1, showing no significant total gene expression outliers compared to whole blood controls from GTEx.

*Supplementary Figure S2: CHD2 protein quantification from induced pluripotent stem cell lines (iPSCs)*

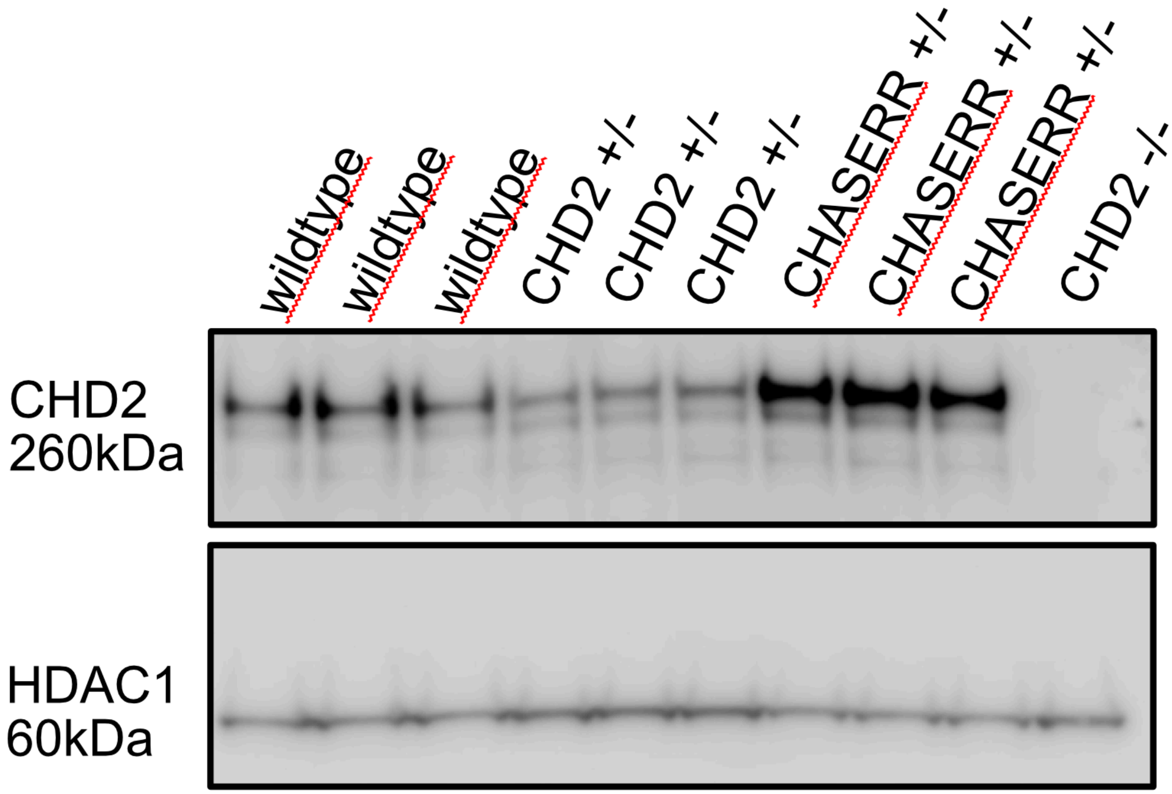


*Supplementary Figure S2 Legend:*

Protein quantification from patient-derived induced pluripotent stem cell (iPSC) lines demonstrates increased CHD2 abundance in Individual 1 (*CHASERR+/-*, three technical replicates from 1 cell line), compared to control lines from three individuals with *CHD2* haploinsufficiency (*CHD2+/-*), three control lines (wildtype), and a *CHD2* knockout iPSC line (*CHD2-/-*).

*Supplementary Methods*

<u>Short-read genome sequencing</u>

For Individual 1, quad genome sequencing was performed for the proband, unaffected sibling, and parents at the Broad Institute Genomics Platform. PCR-free preparation of sample DNA (350 ng input at >2 ng/μl) was accomplished with Illumina HiSeq X Ten v2 chemistry, and libraries were sequenced to a mean coverage of 30x. Genome sequence data were processed through a pipeline based on Picard with base quality score recalibration and local realignment at known indels. The BWA aligner mapped reads to the human genome build 38 (GRCh38). Single nucleotide variants (SNVs) and insertions/deletions (indels) are called jointly across all samples with the Genome Analysis Toolkit (GATK) HaplotypeCaller package version 4.0. Default filters were applied to SNV and indel calls by the GATK Variant Quality Score Recalibration (VQSR) approach. Structural variants (SV) were called using Manta 1.6.0 (Illumina)[1]. Short tandem repeats (STRs) were genotyped by ExpansionHunter[2].

For Individual 2, trio genome sequencing was performed at the French National Research Centre for Human Genomics (CNRGH, Évry, France). Library generation was performed with TruSeq® DNA PCR-free kit, and sequenced on a HiSeq X5 (Illumina). DNA sequences were mapped to the reference human genome build 37 (GRCh37) using bwa 0.7.12 and SNVs and indels were called using DRAGEN Germline 3.8.4 (Illumina). SVs and STRs, were detected using a custom pipeline (https://gitlab.univ-nantes.fr/kriquin/sv-genome). SV are called by Manta 1.6.0 (Illumina)[1] and DELLY 0.8.7 (European Molecular Biology Laboratory)[3] and annotated using AnnotSV 2.2[4]. STRs are genotyped by ExpansionHunter Denovo 0.9.0 (Illumina)[5]. All variants were lifted over and reported here on GRCh38.

For Individual 3, trio genome sequencing was performed AURAGEN laboratory, as part of the French Genomic Initiative PFMG2025. DNA was extracted according to standard procedures and fragmented with Covaris L220plus®. Library generation was performed using TruSeq® DNA PCR-Free kit, (Illumina®, San Diego, CA, USA). Paired-end 2x150bp genome sequencing was performed on a NovaSeq 6000 instrument (Illumina). Reads were aligned to human reference GRCh38.p13 using BWA-MEM 0.7.17, CNVnator v0.4.1 and Manta. SNVs were called using GATK HaplotypeCaller 4.1.8.0 and annotated with Variant Effect Predictor 98.3. Mean depth of sequencing was 43.4x, 47.6x and 49.9x for the proband, mother and father respectively.

<u>Long-read genome sequencing</u>

Long-read sequencing was performed on Individual 1 and parents using the Pacific Biosciences (PacBio) circular consensus sequencing (CCS) protocol. Briefly, for library preparation, 5 μg of high molecular weight genomic DNA (>50% of fragments ≥40 kb) was sheared to ~10 kb using the Megaruptor 3 (B06010003; Diagenode), followed by DNA repair and ligation of PacBio adapters using the SMRTbell Template Prep Kit v1.0 (100-991-900). Libraries were then size selected for 10 kb ± 20% using the SageELF with 0.75% agarose cassettes (Sage Science). Following quantification with the Qubit dsDNA High Sensitivity Assay Kit (Q32854; Thermo Fisher Scientific), libraries were diluted to 50 pM per single molecule, real-time (SMRT) cell, hybridized with PacBio v2 sequencing primer, and bound with SMRT sequencing polymerase using Sequel II Binding Kit 1.0 (101-731-100). CCS sequencing was performed on the Sequel II instrument using 8M SMRT Cells (101-389-001) and Sequel II Sequencing 1.0 Kit (101-717-200), with a 2-h pre-extension time and 30-h movie time per SMRT cell. Initial quality filtering, base calling, and adapter marking were performed automatically on board the Sequel II to generate a BAM file.

Patient-derived cell lines

Patient fibroblasts were generated from Individuals 1 and 2. As a haploinsufficiency control, fibroblasts were generated from an affected individual with a *CHD2* loss of function variant (p.G491VfsX13)[7]. We reprogrammed induced pluripotent stem cells (iPSCs) from fibroblasts for individual 1 and the *CHD2* haploinsufficiency control using the CytoTune-iPS 2.0 Sendai Reprogramming Kit (A16517). A *CHD2*$^{-/-}$ (knockout) line was generated using CRISPR/Cas9 genome editing with sgRNAs targeting exon 5 of *CHD2*. iPSCs were karyotyped using the KaryoStat Assay (Thermo #905403) and assessed for pluripotency markers. These iPSCs were maintained on matrigel (Corning #354277) in mTeSR+ (STEMCELL technologies #100-0276) and differentiated to neural precursor cells (NPCs) using the STEMDiff Neural Induction Medium (STEMCELL Technologies #05835).

RNA and cDNA amplicon sequencing

### Individual 1

RNA from whole blood, skin fibroblasts, iPSCs and NPCs were quantified and processed using a stranded, polyA-tailed kit (Illumina) before being multiplexed, then underwent 150 bp paired-end sequencing with approximately 30-50 million reads generated per sample. The sequencing data were processed with a pipeline adapted from one developed by the GTEx Consortium[8]. Briefly, FASTQ files were aligned to the GRCh38 reference sequence using STAR-2.6.1b in 2-pass mode, and duplicates were marked with Picard. RNA-seq data were de-multiplexed and each sample sequence data aggregated into a single Picard BAM file. We quantified gene expression using RSEM to generate TPM values for expressed genes in each sample. The processed alignment files were then used as input for the outlier detection step.

We isolated RNA from biological replicates of patient-derived fibroblasts, iPSCs and NPCs and performed targeted resequencing of the c.-367G>C 5'UTR variant. Briefly, 1µg of RNA was converted to cDNA using the SuperScript IV Reverse Transcriptase (ThermoFisher #18091200) and the 5'UTR variant containing region was amplified with forward (TCGTCGGCAGCGTCAGATGTGTATAAGAGACA*GTAGCCCCCTGCCTTTCCTA*G) and reverse (GTCTCGTGGGCTCGGAGATGTGTATAAGAGACAG*AAAAATGGAGGTGCGCCATG*) primers. Products were barcoded and sequenced on an Illumina Miniseq, using a 150bp paired end protocol and mapped to the genome (hg38) using BWA, and individual reads containing reference and alternate allele were counted. We also applied this allelic counting method for all single nucleotide variants in *CHD2* (c.-367G>C, rs12906163, rs9672839, rs145018235, rs13759) from RNA-seq data.

### Individual 2

Whole blood samples were collected in PAXgene Blood RNA Tube (BD Biosciences), and fibroblasts were obtained by skin biopsy and cultured by standard techniques. Total RNA from whole blood was extracted with PAXgene Blood RNA Kit (PreAnalytiX), and total RNA from fibroblasts was extracted with NucleoSpin RNA Plus kit (Macherey-Nagel) according to the manufacturer's instructions. mRNA was isolated using NEBNext Poly(A) mRNA Magnetic Isolation Module (New England Biolabs) and libraries prepared using the NEBNext Ultra II DNA Library Prep Kit for Illumina (New England Biolabs). Library quality controls were assessed on TapeStation (4200, High Sensitivity RNA ScreenTape, Agilent), and nucleic concentration of each sample was evaluated on Qubit Fluorometer using Qubit dsDNA High Sensitivity (HS) Assay Kits (Thermo Fisher). Sequencing of 75 bp paired-end reads has been performed on NextSeq 550 Sequencing System (Illumina) using NextSeq 500/550 High Output Kit v2.5 (150 cycles). Approximately 50 million reads were generated per sample. The sequencing data were aligned to the GRCh37/hg19 reference sequence using STAR-2.5.3a. Gene expression was quantified using Kallisto-0.46.2 (Bray et al., 2016) to generate TPM values for expressed genes in each sample. The allelic ratios of rs4777755, rs11074121, and rs2272457 were also quantified from the RNA-seq data.

For *CHD2* allelic imbalance analysis, haplotype-level gene expression from the GTEx Consortium[8] was quantified using phASER[6].

<u>Immunoblot for CHD2 abundance</u>

Western blot analysis was performed in iPSCs in each cell line in biological triplicate using a custom generated CHD2 antibody (Pocono) to 1250-1350 amino acids based on the discontinued CHD2 antibody (abcam #68301). Nuclei are isolated via fractionation and centrifugation using the Abcam Nuclear Fractionation Protocol. Primary antibodies (CHD2, 1:1000) and HDAC1 Mouse mAb (CST #5356, 1:1000) were incubated overnight at 4$^{o}$C in blocking buffer. Membranes were washed 3x 5 minutes in 0.05% PBS-Tween prior to incubation with secondary antibody, anti-rabbit HRP (Abcam #205718, 1:5000) and anti-mouse HRP (Abcam #7076, 1:5000) for 60 minutes at room temperature. After washing 3x 5 minutes in PBS-T, membranes were incubated in Amersham ECL Prime (GE Healthcare) according to manufacturer's instructions. Membranes were imaged on a Licor Odyssey Fc.
